# Effects of Aerobic Exercise on Inflammatory Biomarkers, Pain Intensity, and Quality of Life in Patients with Non-Specific Chronic Low Back Pain: A Randomized Controlled Trial

**DOI:** 10.64898/2026.06.21.26356027

**Authors:** Nweke Vincent Chinonso, Kadiree Ekundayo Fatai, Anelechi Kenneth Madume, Chidiebele P Ojukwu, Adaeze Imelda Onyekwelu, Nweke Queeneth kadilobari, Nweke Augustine Chidera, Charles I. Ezema

## Abstract

**Background:** Non-specific chronic low back pain (NSCLBP) is a major cause of disability worldwide and is associated with low-grade systemic inflammation. This study investigated the effects of aerobic exercise on inflammatory biomarkers, pain intensity, and quality of life among individuals with NSCLBP.

**Methods:** In this parallel-group randomized controlled trial, 41 participants with NSCLBP were allocated to either an aerobic exercise plus health education group (n=21) or a health education-only control group (n=20). Participants in the intervention group completed supervised aerobic cycling three times weekly for 12 weeks. Outcome assessors and laboratory personnel were blinded to group allocation. Outcomes were measured at baseline, Week 8, and Week 12.

**Results:** Significant Time × Group interaction effects were observed for TNF-α (p=0.046), IL-6 (p<0.001), hs-CRP (p<0.001), and pain intensity (p<0.001). Significant improvements were also observed across all WHOQOL-BREF quality-of-life domains (all p<0.05). After adjustment for baseline values and age, participants in the intervention group had significantly lower Week 12 IL-6 (p=0.013), hs-CRP (p<0.001), and pain intensity (p<0.001) than controls. No serious adverse events were reported.

**Conclusions:** Aerobic exercise combined with health education produced greater improvements in inflammatory biomarkers, pain intensity, and quality of life than health education alone among individuals with NSCLBP. These findings support the integration of structured aerobic exercise into rehabilitation programmes for chronic low back pain.

**SUMMARY BOX:** *What is already known on this topic:*

- Non-specific chronic low back pain (NSCLBP) is associated with persistent pain, reduced quality of life, and low-grade systemic inflammation.
- Aerobic exercise is recommended for NSCLBP management and can improve pain and physical function.
- The effects of aerobic exercise on inflammatory biomarkers in individuals with NSCLBP remain incompletely understood.

*What this study adds:*

- A 12-week supervised aerobic exercise programme significantly improved IL-6, hs-CRP, pain intensity, and quality of life compared with health education alone.
- The beneficial effects of aerobic exercise remained significant after adjustment for baseline differences and age.
- No serious adverse events were reported during the intervention period.

*How this study might affect research, practice or policy:*

- Structured aerobic exercise may be incorporated into routine rehabilitation programmes for individuals with NSCLBP.
- The findings support the use of accessible, low-cost exercise interventions in low-resource settings.
- Future studies should investigate the long-term sustainability and mechanisms of exercise-induced improvements.

## Background

Low back pain (LBP) is one of the most prevalent musculoskeletal disorders worldwide and remains the leading cause of years lived with disability (YLDs) [1]. Approximately 85–90% of cases are classified as non-specific, meaning that symptoms cannot be attributed to a specific pathological cause such as fracture, infection, malignancy, or inflammatory disease [2,3]. Chronic low back pain (CLBP), defined as pain persisting for more than 12 weeks, contributes substantially to disability, healthcare utilization, reduced productivity, and impaired quality of life [2]. In low- and middle-income countries, including Nigeria, the burden of chronic musculoskeletal disorders continues to increase because of demographic and lifestyle changes [1].

Growing evidence suggests that non-specific chronic low back pain (NSCLBP) involves biological, psychological, and social factors [2]. Among the biological mechanisms implicated in chronic pain persistence, systemic inflammation has received increasing attention. Elevated concentrations of pro-inflammatory biomarkers, including tumour necrosis factor-alpha (TNF-α), interleukin-6 (IL-6), and C-reactive protein (CRP), have been reported among individuals with NSCLBP and have been associated with greater pain severity, disability, and functional limitation [4,5]. Persistent inflammation may contribute to nociceptor sensitization and altered pain processing, thereby sustaining symptoms beyond the initial tissue insult [4].

Pain intensity is a key clinical outcome in NSCLBP because it affects physical functioning, participation in daily activities, and overall well-being [2]. Persistent pain may also contribute to reduced physical activity, deconditioning, and diminished health-related quality of life [2,6]. Consequently, effective interventions should address both symptom reduction and broader patient-centred outcomes.

Exercise therapy is recommended as a first-line non-pharmacological intervention for chronic low back pain because of its effectiveness, safety, and cost-efficiency [7,8]. Aerobic exercise has attracted particular interest because it may improve physical function, reduce pain, and modulate inflammatory processes simultaneously [9,10]. Regular aerobic exercise has been associated with reductions in circulating inflammatory biomarkers, improved immune regulation, and enhanced quality of life among individuals with chronic musculoskeletal conditions [9–11].

Despite growing evidence supporting aerobic exercise, important knowledge gaps remain. Few studies have simultaneously examined inflammatory biomarkers and patient-reported outcomes such as pain intensity and quality of life in individuals with NSCLBP [4,5]. Furthermore, most evidence has been generated outside Sub-Saharan Africa, limiting its applicability to populations with different environmental, occupational, and healthcare contexts [1,12]. In Nigeria, research has focused largely on prevalence and disability, with limited attention to inflammatory pathways and exercise-mediated physiological adaptations [12].

Therefore, this randomized controlled trial aimed to evaluate the effects and safety of a 12-week aerobic exercise programme on inflammatory biomarkers, pain intensity, quality of life, and adverse events among individuals with NSCLBP in Nigeria. Specifically, the study examined changes in serum TNF-α, IL-6, and CRP concentrations alongside measures of pain intensity and health-related quality of life following participation in a structured aerobic exercise intervention.

## Methods

### Study Design

This study employed a parallel-group randomized controlled trial design to investigate the effects of aerobic exercise on systemic inflammation, pain intensity, and quality of life among individuals with non-specific chronic low back pain (NSCLBP). Participants were randomly assigned to either an aerobic exercise plus health education group or a health education-only control group. Outcome assessments were conducted at baseline (Week 0), Week 8, and Week 12.

The primary outcomes were serum concentrations of tumour necrosis factor-alpha (TNF-α), interleukin-6 (IL-6), and high-sensitivity C-reactive protein (hs-CRP). Secondary outcomes included pain intensity and health-related quality of life. Inflammatory biomarkers and pain intensity were assessed at all three time points, whereas quality-of-life outcomes were assessed at baseline and Week 12. The study was conducted and reported in accordance with the Consolidated Standards of Reporting Trials (CONSORT) 2010 Statement [13,14]. This trial was registered retrospectively with the Pan African Clinical Trials Registry (PACTR; Registration Number: PACTR202606887164888) on 11 June 2026. Registration occurred after participant recruitment had commenced because of administrative delays. The study protocol, eligibility criteria, outcome measures, intervention procedures, and statistical analysis plan were established before recruitment and remained unchanged following registration.

### Study Setting

The study was conducted at the Physiotherapy Department of Rivers State University Teaching Hospital (RSUTH), Port Harcourt, Nigeria. Participant recruitment, intervention delivery, and outcome assessments were conducted within the department and affiliated diagnostic laboratories.

### Sample Size Determination

Sample size was calculated a priori using G*Power version 3.1 [15] for a mixed-design repeated-measures ANOVA. Based on a moderate effect size (f = 0.25), α = 0.05, and 80% power, a minimum sample of 36 participants was required. Allowing for 20% attrition, the target sample size was increased to 44 participants. A total of 41 participants completed the study.

### Interim Analyses and Stopping Guidelines

No interim analyses or formal stopping guidelines were planned because of the short duration and low-risk nature of the intervention.

### Participant Recruitment and Screening

Participants were recruited consecutively from the Physiotherapy Department of RSUTH through referrals from physiotherapists and physicians managing chronic low back pain. Eligibility was determined through medical history review, physical examination, and assessment against predefined eligibility criteria. Written informed consent was obtained from all participants before enrolment. Participant flow is presented in the CONSORT diagram (Figure 1).

**Figure 1.**
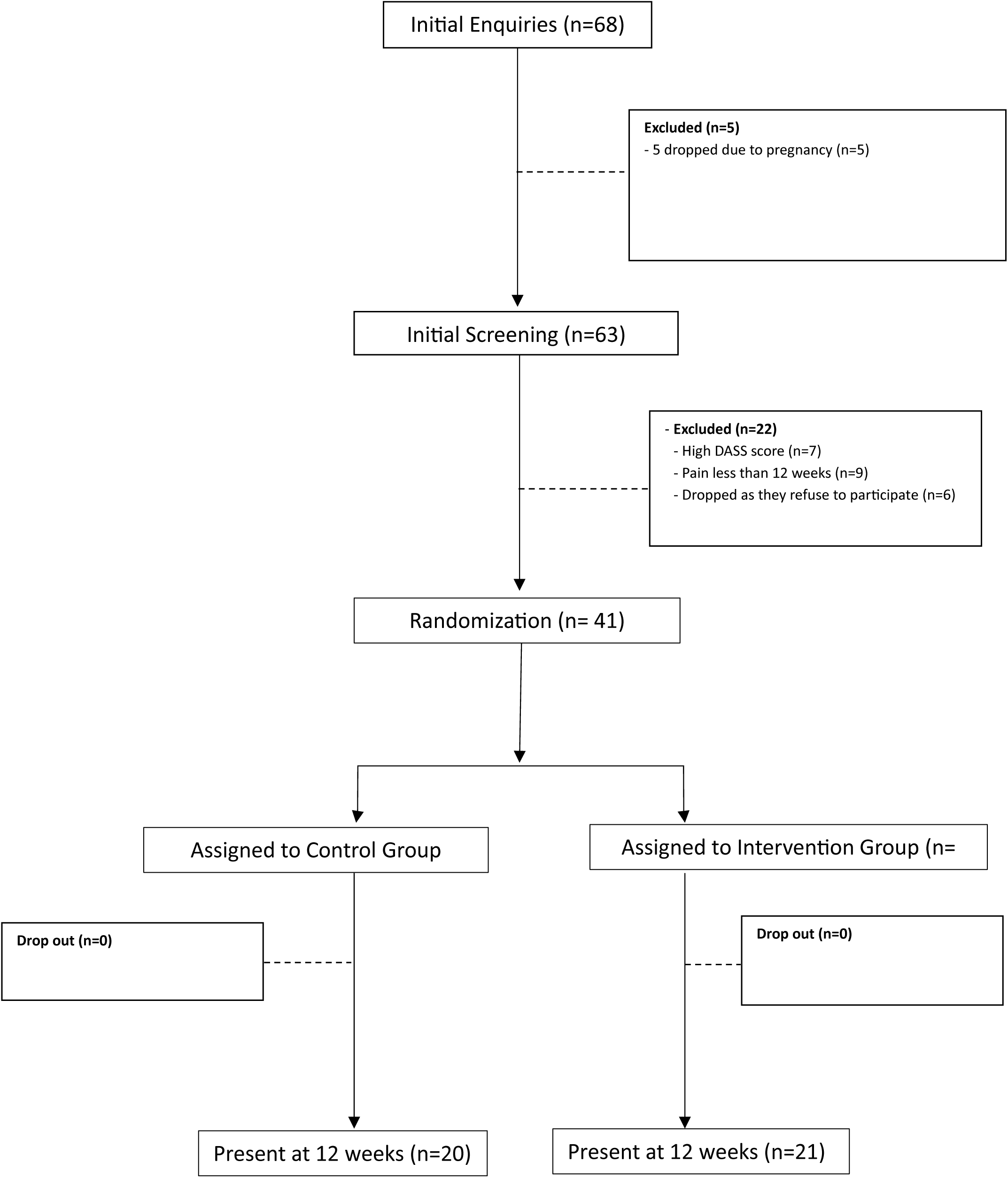
CONSORT Flow Diagram of Participant Recruitment, Randomization, Allocation, Follow-up, and Analysis.

### Eligibility Criteria

#### Inclusion Criteria

Participants were eligible for inclusion if they were aged 18 years or older; had a clinical diagnosis of non-specific chronic low back pain persisting for at least 12 weeks; were medically stable and able to participate in moderate-intensity aerobic exercise; were willing to comply with all study procedures; and provided written informed consent prior to participation.

#### Exclusion Criteria

Participants were excluded if they had a history of spinal surgery; presented with signs of lumbar radiculopathy, spinal stenosis, nerve root compression, or other specific spinal pathologies; had inflammatory rheumatic disorders, spinal infection, vertebral fracture, malignancy, or other systemic inflammatory conditions; were pregnant; demonstrated cognitive impairment that could limit participation or compliance with study procedures; or had uncontrolled cardiovascular, respiratory, neurological, or metabolic disorders that contraindicated participation in aerobic exercise.

#### Eligibility Criteria for Intervention Providers

The aerobic exercise intervention was delivered by licensed physiotherapists with clinical experience in musculoskeletal rehabilitation. All intervention providers received orientation regarding the study protocol prior to participant enrolment.

### Equity, Diversity and Inclusion Statement

The research team considered equity, diversity, and inclusion throughout the study design and conduct. Eligibility criteria were based on clinical characteristics rather than sex, ethnicity, religion, socioeconomic status, or other demographic factors. Both male and female participants were eligible for recruitment. Participants were recruited consecutively from a routine clinical population attending a tertiary healthcare facility, and no groups were intentionally excluded from participation beyond the predefined clinical eligibility criteria.

### Randomization and Allocation Concealment

Following baseline assessment, participants were randomly allocated to either the intervention group or the control group using a computer-generated block randomization sequence with a block size of four and an allocation ratio of 1:1. The random allocation sequence was generated by an independent researcher using a computerized random-number generator. The independent researcher was not involved in participant recruitment, enrolment, intervention delivery, outcome assessment, or data analysis.

Allocation concealment was maintained using sequentially numbered, sealed, opaque envelopes containing group assignments. The envelopes were prepared by the independent researcher and opened only after completion of baseline assessment, thereby preventing foreknowledge of treatment allocation and minimizing selection bias.

### Blinding

Due to the nature of the intervention, blinding of participants and treating physiotherapists was not feasible. However, laboratory personnel responsible for analysing blood samples and research assistants involved in outcome assessment were blinded to group allocation.

### Aerobic Exercise Group

Participants allocated to the intervention group completed a supervised aerobic exercise programme using a Monark Ergomedic 828E cycle ergometer (Monark Exercise AB, Vansbro, Sweden). The programme was prescribed according to the Frequency, Intensity, Time, and Type (FITT) principle and established exercise training guidelines [16,17]. Exercise sessions were conducted three times weekly for 12 weeks under the supervision of a physiotherapist. Intensity was prescribed using Heart Rate Reserve (HRR), calculated using the Karvonen formula [18], and monitored using Borg’s Rating of Perceived Exertion (RPE) Scale [19].

Each session consisted of a 5–10-minute warm-up of low-intensity cycling and flexibility exercises, followed by 30–45 minutes of aerobic cycling. Exercise intensity progressed from 40–50% HRR during Weeks 1–2 to 60–70% HRR by Week 6 and was maintained thereafter. Sessions concluded with 10–15 minutes of low-intensity cycling, stretching, and relaxation exercises. Attendance and adherence were monitored throughout the intervention period.

### Control Group

Participants assigned to the control group received health education on the nature and management of non-specific chronic low back pain, including posture, activity modification, back care principles, and self-management strategies. Participants were instructed to maintain their usual activities and refrain from initiating structured exercise programmes during the study period. Following study completion, control participants were offered physiotherapy management.

### Outcome Measures

Outcome assessments were conducted at baseline (Week 0), Week 8, and Week 12.

### Inflammatory Biomarkers

The primary biological outcomes were serum concentrations of tumour necrosis factor-alpha (TNF-α), interleukin-6 (IL-6), and high-sensitivity C-reactive protein (hs-CRP). Venous blood samples were collected by trained laboratory personnel under standardised conditions at least 48 hours after the most recent exercise session. Biomarker concentrations were measured using commercially available high-sensitivity enzyme-linked immunosorbent assay (ELISA) kits according to manufacturers’ instructions. Laboratory personnel remained blinded to group allocation.

### Pain Intensity

Pain intensity was assessed using the Visual Analogue Scale (VAS), a valid and reliable measure of pain severity in individuals with chronic pain [20].

### Quality of Life

Health-related quality of life was assessed using the World Health Organization Quality of Life Brief Questionnaire (WHOQOL-BREF), which evaluates physical, psychological, social, and environmental domains of quality of life [21].

### Harms Assessment

Adverse events were monitored systematically throughout the intervention period. An adverse event was defined as any undesirable medical occurrence, symptom, injury, or health-related complaint arising during study participation, irrespective of whether it was considered related to the intervention. Participants were asked to report any adverse events during exercise sessions and follow-up assessments. Physiotherapists supervising the intervention documented all reported events, including musculoskeletal discomfort, dizziness, fatigue, cardiovascular symptoms, or any other unexpected occurrence. The severity and potential relationship to the intervention were recorded and reviewed by the research team.

### Ethical Considerations

Ethical approval for the study was obtained from the Rivers State University Teaching Hospital Health Research Ethics Committee (Approval No.: RSUTH/REC/2025897). Administrative permission to conduct the study was obtained from Rivers State University Teaching Hospital. Written informed consent was obtained from all participants prior to enrolment. The study was conducted in accordance with the ethical principles outlined in the Declaration of Helsinki.

### Statistical Analysis

Data were analysed using IBM SPSS Statistics version 24.0 (IBM Corp., Armonk, NY, USA). All randomized participants completed follow-up assessments and were included in the analyses according to the intention-to-treat principle. As there were no missing outcome data, no imputation methods were required. Descriptive statistics were computed for all study variables. Continuous variables were summarised using means and standard deviations (SD), while categorical variables were presented as frequencies and percentages. Data normality was assessed using the Shapiro–Wilk test, histograms, and Q–Q plots. Baseline differences between groups were examined using independent-samples t-tests, Mann–Whitney U tests, Chi-square tests, or Fisher’s exact tests, as appropriate.

Changes in inflammatory biomarkers (TNF-α, IL-6, and hs-CRP) and pain intensity across baseline, Week 8, and Week 12 were analysed using mixed-design repeated-measures analysis of variance (ANOVA), with time as the within-subject factor and treatment group as the between-subject factor. Quality-of-life outcomes, assessed at baseline and Week 12, were analysed using mixed-design repeated-measures ANOVA. Mauchly’s test was used to assess the assumption of sphericity, and Greenhouse–Geisser corrections were applied where violations occurred. Significant main and interaction effects were further explored using Bonferroni-adjusted pairwise comparisons.

Because significant baseline differences were observed for age and IL-6 concentrations, analyses of covariance (ANCOVA) were performed to compare Week 12 IL-6, hs-CRP, and pain intensity outcomes between groups while adjusting for baseline values and age. Estimated marginal means, adjusted mean differences, and 95% confidence intervals were reported where appropriate. Effect sizes were expressed as partial eta squared (ηp²), with values of 0.01, 0.06, and 0.14 interpreted as small, medium, and large effects, respectively. Statistical significance was set at p < 0.05.

## Results

### Participant Flow

Participant recruitment, allocation, follow-up, and analysis are presented in the CONSORT flow diagram (Figure 1). A total of 44 individuals were assessed for eligibility, of whom three were excluded prior to randomization. The remaining 41 participants met the eligibility criteria and were randomly allocated to either the aerobic exercise plus health education group (n = 21) or the health education-only control group (n = 20). No participants were lost to follow-up or withdrew during the study period. Consequently, all randomized participants completed the 12-week intervention and were included in the final analyses.

### Adverse Events

No serious adverse events, exercise-related injuries, or unintended effects were reported during the intervention period in either group.

### Participant Characteristics at Baseline

Baseline characteristics are presented in Table 1. Groups were generally comparable at baseline. However, participants in the control group were significantly older than those in the intervention group (p=0.037), while baseline IL-6 concentrations were significantly higher in the intervention group (p<0.001). No other significant between-group differences were observed.

**Table 1.** Participant Characteristics at Baseline.

| Variable | Intervention (n = 21) | Control (n = 20) | p-value | Effect Size |
| --- | --- | --- | --- | --- |
| Age (years) | 35.19 ± 7.92 | 39.55 ± 4.50 | 0.037 | d = 0.68 |
| Sex, n (%) |  |  | 0.853 | V = 0.03 |
| Male | 12 (57.1) | 12 (60.0) | – | – |
| Female | 9 (42.9) | 8 (40.0) | – | – |
| Occupation, n (%) |  |  | 0.239 | V = 0.44 |
| Civil Servant | 5 (23.8) | 10 (50.0) | – | – |
| Trader | 6 (28.6) | 4 (20.0) | – | – |
| Rig Worker | 4 (19.0) | 1 (5.0) | — | — |
| Health Worker | 4 (19.0) | 3 (15.0) | — | — |
| Farmer | 0 (0.0) | 2 (10.0) | — | — |
| Police Officer | 1 (4.8) | 0 (0.0) | — | — |
| Student | 1 (4.8) | 0 (0.0) | — | — |
| Height (m) | 1.62 ± 0.09 | 1.66 ± 0.07 | 0.126 | d = 0.49 |
| Weight (kg) | 80.80 ± 13.13 | 82.85 ± 16.22 | 0.657 | d = 0.14 |
| BMI (kg/m<sup>2</sup>) | 31.02 ± 6.35 | 30.05 ± 6.04 | 0.621 | d = 0.16 |
| Duration of Pain (months) | 12.00 ± 2.05 | 12.50 ± 0.95 | 0.321* | d = 0.31 |
| TNF-α (pg/mL) | 18.25 ± 2.52 | 17.66 ± 3.00 | 0.166 | r = 0.22 |
| IL-6 (pg/mL) | 6.96 ± 1.43 | 4.37 ± 1.66 | <0.001 | d = 1.67 |
| hs-CRP (mg/L) | 7.07 ± 2.60 | 6.23 ± 1.66 | 0.226 | d = 0.38 |
| Pain Intensity (VAS) | 8.05 ± 1.32 | 8.75 ± 2.36 | 0.491 | r = 0.11 |

Note: Data are presented as mean ± SD unless otherwise stated. BMI = body mass index; TNF-α = tumour necrosis factor-alpha; IL-6 = interleukin-6; hs-CRP = high-sensitivity C-reactive protein; VAS = visual analogue scale.

### Effects of Aerobic Exercise on Inflammatory Biomarkers

Changes in inflammatory biomarkers are presented in Table 2. Significant Time × Group interaction effects were observed for TNF-α (p=0.046), IL-6 (p<0.001), and hs-CRP (p<0.001), indicating greater improvements in the intervention group compared with the control group. The largest effects were observed for hs-CRP (ηp²=0.428) and IL-6 (ηp²=0.364). Detailed descriptive statistics and ANOVA results are shown in Table 2.

**Table 2.**
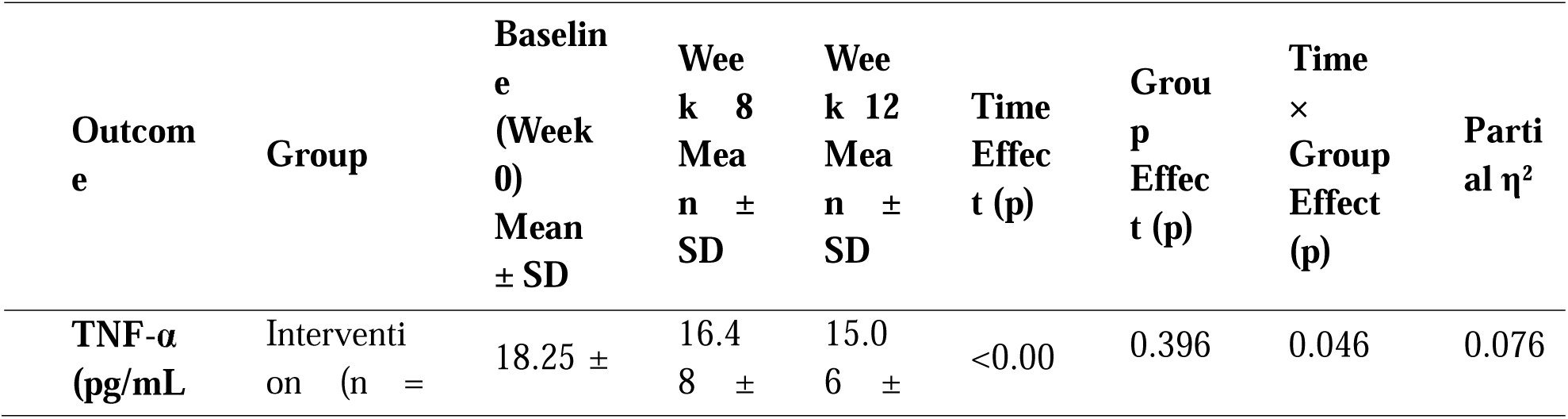
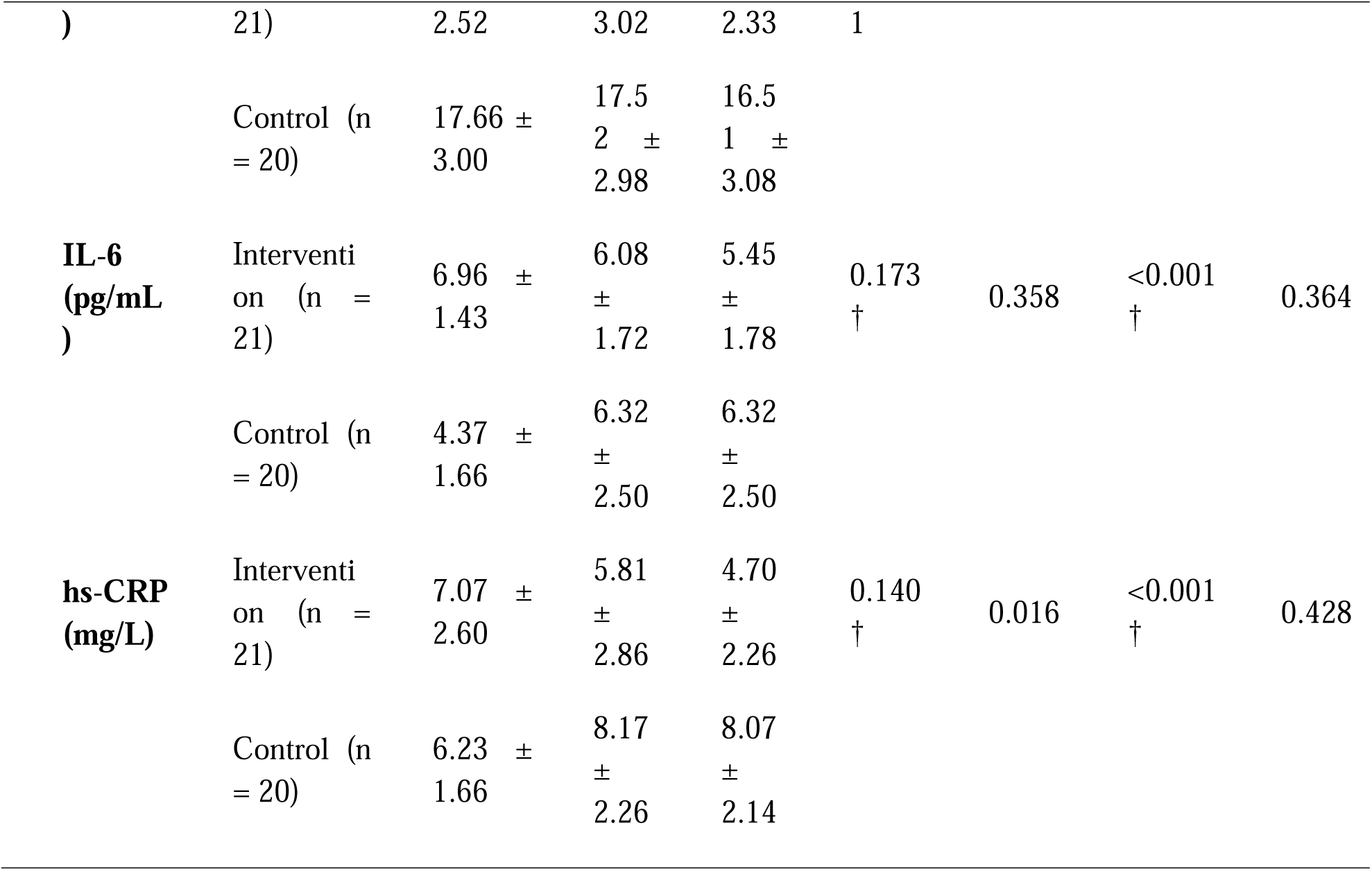
Effects of Aerobic Exercise on Inflammatory Biomarkers (TNF-α, IL-6, and hs-CRP)

### Effects of Aerobic Exercise on Pain Intensity and Quality of Life

Pain intensity and quality-of-life outcomes are presented in Table 3. Significant effects of time, group, and Time × Group interaction were observed for pain intensity (all p<0.001), indicating greater reductions in pain among participants receiving aerobic exercise. Significant Time × Group interaction effects were also observed across all WHOQOL-BREF domains (all p≤0.001), demonstrating greater improvements in physical, psychological, social, and environmental quality of life in the intervention group compared with the control group.

**Table 3.** Effects of Aerobic Exercise on Pain Intensity and Quality of Life.

| Outcome | Group | Baseline<br>Mean<br>± SD | Post-Intervention<br>Mean<br>± SD | Time<br>Effect<br>(p) | Group<br>Effect<br>(p) | Time<br>×<br>Group<br>Effect<br>(p) | Partial $\eta^2$ |
| --- | --- | --- | --- | --- | --- | --- | --- |
| Pain Intensity<br>(VAS) | Intervention (n = 21) | 8.05 ± 1.32 | 3.57 ± 1.03 | <0.001 | <0.001 | <0.001 | 0.213 |
|  | Control (n = 20) | 8.75 ± 2.36 | 6.70 ± 1.13 |  |  |  |  |

| Outcome | Group | Baseline Mean ± SD | Post-Intervention Mean ± SD | Time Effect (p) | Group Effect (p) | Time × Group Effect (p) | Partial $\eta^2$ |
| --- | --- | --- | --- | --- | --- | --- | --- |
| Physical QoL | Intervention (n = 21) | 2.98 ± 0.25 | 3.41 ± 0.34 | 0.001 | 0.006 | 0.001 | 0.266 |
| | Control (n = 20) | 3.02 ± 0.23 | 3.00 ± 0.28 | | | | |
| Psychological QoL | Intervention (n = 21) | 2.97 ± 0.49 | 3.63 ± 0.54 | 0.003 | 0.015 | 0.001 | 0.234 |
| | Control (n = 20) | 3.03 ± 0.46 | 3.00 ± 0.43 | | | | |
| Social QoL | Intervention (n = 21) | 2.62 ± 0.74 | 3.62 ± 0.80 | 0.003 | 0.015 | 0.001 | 0.234 |
| | Control (n = 20) | 2.72 ± 0.69 | 2.67 ± 0.65 | | | | |
| Environmental QoL | Intervention (n = 21) | 2.83 ± 0.74 | 3.83 ± 0.80 | 0.003 | 0.015 | 0.001 | 0.234 |
| | Control (n = 20) | 2.93 ± 0.69 | 2.88 ± 0.65 | | | | |

### Adjusted Between-Group Comparisons at Week 12

ANCOVA was performed to adjust for baseline differences in age and outcome values. After adjustment, participants in the intervention group demonstrated significantly lower IL-6 (p=0.013), hs-CRP (p<0.001), and pain intensity (p<0.001) at Week 12 compared with the control group. The largest adjusted effect was observed for pain intensity (ηp²=0.698), followed by hs-CRP (ηp²=0.516) and IL-6 (ηp²=0.157). Adjusted estimates are presented in Table 4.

**Table 4.** Adjusted Between-Group Comparisons at Week 12 (ANCOVA)

| Outcome | Adjusted Mean Intervention (95% CI) | Adjusted Mean Control (95% CI) | Adjusted Mean Difference (95% CI) | F (df) | p-value | Partial $\eta^2$ |
| --- | --- | --- | --- | --- | --- | --- |
| IL-6 (pg/mL) | 4.72 (3.61–5.82) | 7.09 (5.95–8.24) | -2.38 (-4.21 to -0.54) | F(1,37)=6.876 | 0.013 | 0.157 |
| hs-CRP (mg/L) | 4.36 (3.48–5.24) | 8.42 (7.52–9.32) | -4.06 (-5.37 to -2.75) | F(1,37)=39.493 | <0.001 | 0.516 |

| **Outcome** | **Adjusted Mean Intervention (95% CI)** | **Adjusted Mean Control (95% CI)** | **Adjusted Mean Difference (95% CI)** | **F (df)** | **p-value** | **Partial $\eta^2$** |
| --- | --- | --- | --- | --- | --- | --- |
| Pain Intensity (VAS) | 3.55 (3.08–4.02) | 6.72 (6.24–7.21) | -3.18 (-3.87 to -2.48) | F(1,37)=85.68<br>2 | <0.001 | 0.698 |

## Discussion

This randomized controlled trial investigated the effects of a 12-week aerobic exercise programme on inflammatory biomarkers, pain intensity, and quality of life among individuals with non-specific chronic low back pain (NSCLBP). Participants who received aerobic exercise in addition to health education demonstrated significantly greater improvements in inflammatory biomarkers, pain intensity, and quality of life than those who received health education alone. Significant Time × Group interaction effects were observed for TNF-α, IL-6, and hs-CRP, while ANCOVA confirmed that improvements in IL-6, hs-CRP, and pain intensity remained significant after adjustment for baseline values and age. These findings support the potential anti-inflammatory and clinical benefits of aerobic exercise for individuals with NSCLBP.

### Effects of Aerobic Exercise on Inflammatory Biomarkers

Participants who received aerobic exercise demonstrated significantly greater reductions in IL-6 and hs-CRP concentrations than those receiving health education alone. These findings are consistent with evidence showing that regular aerobic exercise may reduce chronic low-grade inflammation through improvements in immune regulation, metabolic health, and anti-inflammatory signalling pathways [11]. Previous reviews have similarly reported reductions in inflammatory biomarkers following exercise interventions among individuals with chronic low back pain [5]. Elevated concentrations of IL-6, CRP, and TNF-α have been associated with greater pain severity and disability in NSCLBP, supporting the role of inflammation in chronic pain persistence [4].

Although TNF-α also improved, the observed effect was smaller than those for IL-6 and hs-CRP. Similar findings have been reported previously, with TNF-α often demonstrating less consistent responses to exercise interventions than other inflammatory biomarkers [5,11]. The observed biomarker changes should be interpreted as physiological adaptations associated with participation in the exercise programme rather than evidence of a direct causal mechanism linking inflammation reduction to improvements in pain or quality of life.

### Effects of Aerobic Exercise on Pain Intensity

Participants who received aerobic exercise experienced significantly greater reductions in pain intensity than those receiving health education alone. These findings are consistent with systematic reviews and clinical practice guidelines recommending exercise therapy as a first-line intervention for chronic low back pain [7,8,24]. Exercise may reduce pain through improvements in physical conditioning, endogenous pain modulation, movement confidence, and reductions in fear-avoidance behaviours [25].

The reduction in pain intensity observed in the intervention group was not only statistically significant but also clinically meaningful. Participants demonstrated an average reduction of approximately 4.5 points on the Visual Analogue Scale, substantially exceeding the minimal clinically important difference reported for chronic musculoskeletal pain [26].

### Effects of Aerobic Exercise on Quality of Life

Significant improvements were observed across all WHOQOL-BREF domains among participants receiving aerobic exercise. These findings are consistent with previous evidence demonstrating that exercise interventions can improve health-related quality of life, physical functioning, and overall well-being among individuals with chronic low back pain and other chronic musculoskeletal conditions [24]. Reduced pain, improved physical conditioning, and greater participation in daily activities may have contributed to the observed improvements across physical, psychological, social, and environmental domains [6,25].

### Clinical Implications

The findings have important implications for clinical practice, particularly in low-resource settings. Aerobic exercise is relatively inexpensive, accessible, and feasible to implement within routine physiotherapy services. Incorporating structured aerobic exercise into rehabilitation programmes may improve both inflammatory status and patient-centred outcomes among individuals with NSCLBP.

### Strengths and Limitations

Strengths of this study include its randomized controlled design, assessment of objective inflammatory biomarkers alongside patient-reported outcomes, repeated outcome measurements, and use of ANCOVA to account for baseline differences. However, significant baseline differences were observed for age and IL-6 despite randomization. Participants were recruited from a single tertiary healthcare facility, which may limit generalisability. The modest sample size may also have reduced statistical power to detect smaller intervention effects, particularly for TNF-α. Finally, the absence of long-term follow-up precludes conclusions regarding the sustainability of treatment effects.

Future studies should employ larger multicentre trials with longer follow-up periods and explore potential mechanisms linking exercise, inflammation, and clinical outcomes.

## Conclusion

Aerobic exercise in addition to health education was associated with significant improvements in inflammatory biomarkers, pain intensity, and quality of life among individuals with non-specific chronic low back pain. These findings support the integration of structured aerobic exercise into routine rehabilitation programmes for chronic low back pain, particularly in low-resource settings where accessible and cost-effective interventions are needed.

## Data Availability

All data produced in the present study are available upon reasonable request to the authors

## DECLARATIONS

### Author Contributions

VN and CE conceived and designed the study. VN, CE, CO, OA, and KE developed the methodology. VN, AM, OA, and KE collected the data. VN, OA, and KE performed the statistical analyses. VN, OA and KE interpreted the data and drafted the manuscript. VN, CO and CE critically revised the manuscript for important intellectual content. All authors reviewed and approved the final manuscript.

### Guarantor

VN accepts full responsibility for the finished work, had access to the data, and controlled the decision to publish.

## Acknowledgements

The authors sincerely acknowledge all participants who volunteered to take part in this study. Appreciation is extended to the physiotherapists, laboratory personnel, and research assistants whose contributions facilitated participant recruitment, intervention delivery, and data collection.

## Ethics Approval

Ethical approval was obtained from the Rivers State University Teaching Hospital Health Research Ethics Committee (Approval No.: RSUTH/REC/2025897).

## Clinical Trial Registration

Pan African Clinical Trials Registry (PACTR), Registration Number: PACTR202606887164888; retrospectively registered on 11 June 2026. Written informed consent was obtained from all participants prior to enrolment.

## Protocol Availability

The study protocol and statistical analysis plan are available from the corresponding author upon reasonable request.

## Patient and Public Involvement

Patients and members of the public were not involved in the design, conduct, reporting, or dissemination plans of this research.

## Data Sharing Statement

The de-identified participant dataset supporting the findings of this study, together with the statistical analysis code and data dictionary, are available from the corresponding author upon reasonable request and subject to institutional ethical approval requirements.

## List of Abbreviations

ANCOVA: Analysis of Covariance
BMI: Body Mass Index
CONSORT: Consolidated Standards of Reporting Trials
ELISA: Enzyme-Linked Immunosorbent Assay
HRR: Heart Rate Reserve
hs-CRP: High-Sensitivity C-Reactive Protein
IL-6: Interleukin-6
NSCLBP: Non-Specific Chronic Low Back Pain
PPI: Patient and Public Involvement
RCT: Randomized Controlled Trial
RPE: Rating of Perceived Exertion
SD: Standard Deviation
TNF-α: Tumour Necrosis Factor-Alpha
VAS: Visual Analogue Scale
WHOQOL-BREF: World Health Organization Quality of Life Brief Questionnaire

